# Multimodal Wearable and Survey Data Reveal Distinct Physiologic Profiles in Hypermobile-Ehlers Danlos Syndrome for Screening Advancements

**DOI:** 10.64898/2026.04.01.26349981

**Authors:** Damen Wilson, Maureen Shilling, Thomas Nowak, John M. Wo, Clair A Francomano, Thomas Everett, Steven Steinhubl, Matthew Ward

**Affiliations:** Biomedical Engineering, Purdue University, West Lafayette, IN; Division of Gastroenterology & Hepatology, Indiana University School of Medicine, IN; Medical and Molecular Genetics, Indiana University School of Medicine, IN; Krannert Cardiology Research Center, Division of Cardiovascular Medicine, Indiana University School of Medicine, IN; School of Medicine, Indiana University, Indianapolis, IN

**Keywords:** autonomic nervous system, hypermobile Ehlers-Danlos syndrome, heart rate variability, wearable sensors, digital biomarker

## Abstract

Hypermobile Ehlers-Danlos Syndrome (hEDS) is a genetic connective tissue disorder characterized by hypermobile joints, chronic pain, fatigue, brain fog, orthostatic intolerance, and GI symptoms and dysmotility. Its heterogeneous presentation contributes to poor quality of life, inappropriate interventions, and prolonged diagnostic delays, often up to 10 years. This study primarily aimed to determine if physiological signals captured by a medical-grade wrist wearable could characterize autonomic patterns in hEDS and relate them to symptoms. Individuals with hEDS (n=30) and healthy controls (n=28) wore a medical grade smartwatch for 30 days, collecting continuous heart rate variability, activity, oxygen saturation, and blood pressure, alongside initial baseline symptom and quality-of-life surveys. Individuals with hEDS showed greater instability and variability in both systolic and diastolic blood pressure as well as the HRV metric LF/HF ratio, in comparison to healthy controls (*p*-values: 0.04, 0.02, 0.02). During sleep, metrics of parasympathetic activity (HRV measures: HF power, pNN50, RMSSD) trended lower in hEDS than healthy in comparison. As expected, survey domains assessing physiologic symptoms and quality-of-life were significantly worse in the hEDS cohort (*p-*values < 0.05). Notably, autonomic metrics correlated with GI symptoms in the hEDS cohort (Spearman’s ρ range: 0.38-0.60), and psychological symptoms in the healthy cohort (Spearman’s ρ range:-0.47-0.41). Principal component analysis (PCA) of physiologic and symptom features clearly separated groups, supporting distinct physiologic profiles. Combination of GI symptom index and wearable monitoring show promise as a hybrid screening approach that could substantially shorten the time to diagnosis in this population.

## INTRODUCTION

### Hypermobile-Ehlers Danlos Syndrome

Hypermobile Ehlers-Danlos Syndrome (hEDS) is a heterogenous inherited connective tissue disorder that is characterized by musculoskeletal symptoms, joint hypermobility, and tissue fragility, that are associated with a wide range of various systemic manifestations (1). It is a subtype of a larger group of connective tissue disorders called Ehlers-Danlos syndromes (EDS) consisting of 13 types. Many types of EDS are known to be caused by gene alterations affecting collagen function and production; however, hEDS has not been linked to a specific gene or collagen issue as many of the other subtypes have, resulting in key questions unanswered on pathophysiology and diagnosis of the condition. It is theorized that connective tissue plays a part, as well as presence of neuropathy of small fibers may both be contributing to the condition and its unique presentations (2,3).

hEDS is considered a rare disease but is the most prevalent subtype of EDS, with overall EDS estimated to be found in 1 in 5000 individuals, and hEDS accounting for 90% of those EDS cases(2). However, it is thought that this prevalence is under representative due to misdiagnoses and underdiagnoses (4,5). The manifestation of hEDS is very diverse and often includes (but not limited to) the following: joint hypermobility, pain, dislocations and subluxations, fatigue, and autonomic dysfunction with gastrointestinal and cardiovascular symptoms(2). Gastrointestinal manifestations include reflux, heartburn, bloating, abdominal pain, irritable bowel syndrome, and diarrhea (2,6).

Cardiovascular manifestations include postural orthostatic tachycardia syndrome (POTS), neurally-mediated hypotension (NMH), and orthostatic intolerance (2).

### Diagnostic Issues

The diagnosis of hEDS is inherently clinical, as there are no definitive genetic diagnostic tests for patients (7,8). Current diagnosis of hEDS is made in an adult based upon criteria of generalized joint hypermobility, evidence of a generalized musculoskeletal or connective tissue disorder, and exclusion of other diagnoses (9). It is heavily dependent on a Beighton Score, a 9 point scoring system for joint hypermobility; however, this is criticized technique that predominantly tests upper limb joints as opposed to others, thus may be contributing to underdiagnosis (10). Notably though, the current diagnostic criteria omit formal consideration of many quality-of-life symptoms such as fatigue, and cardiovascular and gastrointestinal disturbances, which also may support why current methods of diagnosis are inadequate and have much room for improvement.

As seen with other clinical based diagnosed conditions, this pure clinical approach often leads to delays in treatments and both missed and incorrect diagnoses. A large multi country survey was done to explore rare diseases and their difficulties in diagnosis and management, of which included EDS(5). It was found that in half of EDS patients, there is a 14 year delay between first symptoms and a correct diagnosis(5). In part this is due to misdiagnosis, where if an individual with EDS was misdiagnosed with a psychological/psychiatric condition their delay in correct diagnosis was observed to be 22 years(5).

These delays in diagnosis and treatments can greatly alter an individual’s quality of life, as its been shown hEDS has a severely strong affect in lowering quality of life (2,11). Among rare diseases, EDS has the highest rate of denial of treatment, in part due to lack of diagnosis (5). Furthermore, EDS had the most reported medical (physical, psychological) and non-medical (family disruptions, loss of trust in medicine) consequences directly due to their delay in diagnosis(5).

### Prior Autonomic Physiology Research

Though exact mechanisms are debated, it is known that individuals with hEDS often have symptoms associated with autonomic dysfunction (12). De Wandele et al. performed autonomic function testing on 39 individuals with hEDS and found that there was significant increased sympathetic activity during their 15-min rest period when observing HRV. Furthermore, they found that cardiovascular and sudomotor dysfunction both were evident in hEDS(13). Sleep is of particular interest with hEDS as a majority of individuals with hEDS report inadequate sleep (14,15). Autonomic function during sleep, to the best of our knowledge, has not been studied in hEDS. Sleep-wake patterns, as well as circadian rhythms, can reveal ample information about autonomic function and health (16). Both are known to affect one’s heartrate and blood pressure in predictable 24hr variations in a healthy individual (16–19). Longer term biometric recordings, tracking one’s typical day-to-day profile, have the potential to be measured with the rise in wearable sensors(20). With the advances in wearable health monitors and smart devices, access to more parameters and frequent intervals of sampling may enable a new frontier in understanding physiology and diagnostics. To our knowledge, only a single study has utilized wearables thus far for longer term recording in individuals with hEDS (21). Mathena et al. utilized wearables over the course of year and found that on days individuals had gastrointestinal symptoms, their HRV was also significantly changed from days with minimal symptoms. These insights are an example of wearable based information integrated for the stratification of hEDS disease symptomatology.

### Study Aim

The objective of this study is to evaluate measures of circadian autonomic function from wearables and investigate their relationship with symptoms. Specifically, this study aimed to leverage medical grade wrist sensor-acquired data over long-term recordings to assess underlying autonomic physiology of hEDS, particularly heartrate variability. Additionally, this study aims to explore the feasibility of using wearable derived metrics to distinguish between individuals with and without hEDS, and to identify candidate metrics for future predictive modeling, enabling a new frontier in diagnostics and screening in this disease space.

## MATERIALS AND METHODS

### Study Participants

Sixty participants were recruited for this study, thirty healthy controls and thirty individuals with hEDS. Participants were recruited from the Medical & Molecular Genetics Clinic and Gastrointestinal Motility Clinic at IU School of Medicine, and outreach via social media, flyers, and email listservs.

Participants in the hEDS cohort were eligible if they were: age 18 to 80, of any sex or race, had a confirmed hEDS diagnosis by a Geneticist or Rheumatologist, able to comply with study measures, and were able to use a smartwatch and associated smartphone application. Participants in the healthy cohort were eligible if they were: age 18 to 80, of any sex or race, had no prior GI or autonomic diagnoses, able to comply with study measures, and were able to use a smartwatch and associated smartphone application. Exclusion criteria for both cohorts were inability to provide consent, currently pregnant, or a prisoner.

### Study Approval

Participants were enrolled from July 2024 to October 2025. The experimental protocol was approved by the Indiana University Human Subjects & Institutional Review Board (No. 2006075899). This trial was registered with ClinicalTrial.gov under NCT06491758-2006075899-B. All participants signed informed consent forms prior to beginning the study.

### Study Support

This study was supported by NIH 1T32DC016853, NIH T32GM148382, and the Ehlers Danlos Society.

### Experimental Protocol

#### Initial Visit

Participants first attended an initial in-clinic onboarding visit, during which eligibility was confirmed through a review of inclusion and exclusion criteria, as well as a detailed medication list. Following enrollment in the initial visit, participants completed four surveys, including: Patient Assessment of Gastrointestinal Disorders - Symptom Severity Index (PAGI-SYM), Patient Assessment of Upper Gastrointestinal Disorders - Quality of Life (PAGI-QOL), Composite Autonomic Symptom Scale 31 (COMPASS 31), and Patient-Reported Outcomes Measurement Information System (PROMIS-29).

Participants were provided with the Corsano Cardio Watch 287-2B bracelet (Corsano Health, Switzerland) and received comprehensive training on its use and care. In-clinic, the research team ensured that all device features such as cuffless blood pressure, heart rate, respiratory rate, oxygen saturation, body temperature, activity tracking, and sleep diaries were activated and functioning correctly. Participants were instructed to download the Corsano app on their personal smartphones (compatible with both iOS and Android platforms). The smartwatch data were automatically transmitted via Bluetooth to a HIPAA-compliant deidentified Corsano Medical Cloud Storage platform (the Corsano study portal). Participants then underwent a cuffed blood pressure calibration measurement to ensure accurate monitoring and compliance with the EU-MDR certified cuffless blood pressure measurements during the study period.

#### Study Period

Participants were instructed to wear the smartwatch continuously for 30 days, removing it only for charging and during showers/aquatic activities. The device was to be worn at all other times, including during sleep. Throughout the study period, participants were instructed to journal any symptoms experienced using the app’s symptom diary feature or personal methodologies. To ensure compliance and device functionality, a clinical coordinator contacted participants via email or phone on days 8, 15, and 22 to address any issues and confirm that symptoms were being journaled as instructed. On day 30, participants were instructed to remove the smartwatch and return it to the study site.

### Biometric Data

The Corsano Cardio Watch 287-2B continuously collected a comprehensive range of biometric data throughout the study period (22,23). The following physiological and activity-related parameters were recorded and securely transmitted to the study portal: photoplethysmography (PPG) and pulse rate, RR intervals, respiratory rate, activity metrics (steps, cadence, speed, and estimates energy expenditure), sleep monitoring (sleep duration, quality, and stages), core body temperature, oxygen saturation (SpO₂), and heart rate variability (HRV) metrics. Blood pressure is determined via pulse-wave analysis, which has been validated against 24-ambulatory standard cuff measurement (24).

The following validated HRV metrics were calculated via Corsano to their web portal on average every 60 seconds (25):

- RR Interval: The time (in milliseconds) between consecutive heartbeats, serving as the basis for all HRV analyses.
- SDNN (Standard Deviation of NN Intervals): Reflects overall HRV by measuring the standard deviation of all normal-to-normal (NN) RR intervals, variation often attributed to parasympathetic driven respiratory sinus arrythmia.(26)
- RMSSD (Root Mean Square of Successive Differences): Indicates short-term components of HRV, sensitive to parasympathetic (vagal) activity. (26)
- pNN50: The percentage of successive RR intervals that differ by more than 50 ms, another marker correlated with parasympathetic activity. (26)
- LF (Low Frequency Power): Represents the power in the low-frequency band (typically 0.04–0.15 Hz), thought to be associated with both sympathetic and parasympathetic modulation. (26,27)
- HF (High Frequency Power): Power in the high-frequency band (0.15–0.40 Hz), primarily thought to reflect parasympathetic activity. (26,27)
- VLF (Very Low Frequency Power): Power in the very low frequency band (<0.04 Hz), the physiological significance of which is less well understood but may relate to inflammation and stress responses. (28)
- LF/HF Ratio: Reflects the relative distribution of power between low-and high-frequency HRV components; however, its physiological interpretation is complex and does not provide a direct or reliable measure of sympathovagal balance.(29)

### Questionnaires

#### PAGI-SYM

The Patient Assessment of Upper Gastrointestinal Disorders-Symptom Severity Index (PAGI-SYM) is a 20-item validated questionnaire designed to measure symptom severity in patients with upper gastrointestinal disorders, including gastroparesis, gastroesophageal reflux disease, and functional dyspepsia. It assesses six subscales: heartburn/regurgitation, postprandial fullness/early satiety, nausea/vomiting, bloating, upper abdominal pain, and lower abdominal pain. Participants rate symptom severity over the preceding two weeks using a 6-point Likert scale, with higher scores indicating greater symptom burden. The instrument has demonstrated strong reliability, validity, and responsiveness in clinical settings, making it a foundation for quantifying gastric symptomatology in research and clinical practice. (30)

#### PAGI-QUOL

The Patient Assessment of Upper Gastrointestinal Disorders-Quality of Life (PAGI-QOL) is a 30-item companion to the PAGI-SYM, evaluating disease specific quality of life impacts across five domains: daily activities, clothing, diet/food habits, relationships, and psychological well-being. Scores are inverse, with higher values reflecting better quality of life. The tool exhibits similar validity with other generic measures of quality of life such as the SF-36. Its development and past validation across diverse cohorts ensure its usability in various populations with upper GI disorders. (31)

#### COMPASS-31

The Composite Autonomic Symptom Score-31 (COMPASS-31) is a 31 item questionnaire assessing autonomic dysfunction across six weighted domains: orthostatic intolerance, vasomotor, secretomotor, gastrointestinal, bladder, and pupillomotor symptoms. Its total score (range: 0–100) provides a quantitative index of autonomic symptom burden, allowing tiering and classification of autonomic disease severity in clinical and research settings. (32)

#### PROMIS-29

The Patient-Reported Outcomes Measurement Information System-29 (PROMIS-29) is a generic health related quality of life tool developed by the NIH, assessing seven domains: physical function, anxiety, depression, fatigue, sleep disturbance, social participation, and pain interference. Each domain uses a 4-week recall period and generates T scores normalized to the general population (mean = 50, SD = 10). Although it is not disease specific, its modular design allows integration with chronic disease research. The PROMIS-29’s validity and reliability have been established across diverse populations. (33)

## Data Analysis

All data analysis was performed using custom scripts and functions in MATLAB (The MathWorks, Natick, MA), including data preprocessing, statistical analyses, and visualization, in accordance with standard practices for computational research. This was a pilot exploratory analysis intended to assess the feasibility of using wearable derived biometrics to distinguish between participants with and without hEDS, as well as learn key insights on their physiology. No formal predictive model was developed due to sample size limitations. Instead, descriptive statistics, group comparisons, and exploratory receiver operating characteristic (ROC) curve analyses were performed to identify potential biometrics/transformations for future investigations.

### Windowing

The metrics mentioned prior were pulled from Corsano’s servers for each individual following completion of the study. Each metric was output on average 1 per minute over the 30 days recording for each individual. First windowing was performed such that beginning at time 00:00 or midnight, data was averaged every 30 minutes, such that 48 data points for each metric would occur roughly every 24-hour cycle. This was typically the case, but sometimes due to noise or not wearing the watch a data point may not have been found. In the event data is missing, it was time stamped and treated as NaN, or blanked out, such that replacement or interpolation was not performed to maintain true data measurements.

Overall, for each metric, windowing resulted in approximately 1440 time stamped data points spaced in 30 minute intervals for each participant.

### Night and Day Assessment

In order to observe nighttime and daytime patterns, averaging was performed on each time point for each participant. For example, if a participant had 30 consecutive LF Power values at 01:30 over the course of 30 days, a single data point for 01:30 was found for this individual by averaging those 30 points. In doing so, an average 24-hour pattern can be visualized that is likely more reflective of an individual’s patterns due to the culmination of 30 days of data. To quantify the difference in day vs. night of metrics the time periods of 01:00 to 03:00 for night and 11:00 to 13:00 for day were selected. For these two time windows all values within those windows were averaged together to allow a representative mean for each of the two states of sleep and wake. This was done for each individual’s 24 hour mean on each metric.

### Correlations

To assess the relationship between individual survey responses and autonomic responses, we performed correlation analyses between participants’ survey scores and their corresponding day night differences for each biometric measure. For each comparison, we used the corr function in MATLAB to compute Pearson correlation coefficients the associated p-value for each test. Prior to analysis, data were assessed for normality using visual inspection of histograms and Shapiro-Wilk tests, and Pearson correlations were applied where assumptions of normality were met. If assumptions were violated, Spearman rank correlations were used instead.

### Receiver Operator Curves

To evaluate the diagnostic ability of wearable derived metrics for distinguishing between participants with and without hEDS, we performed receiver operating characteristic (ROC) curve analyses. For each average physiological metric across their month interval analyzed, a ROC curve was generated by plotting the true positive rate against the false positive rate, or at different threshold levels. The area under the ROC curve (AUC) was computed for each metric to quantify overall classification ability. An AUC of 0.5 has traditionally been indicated as having no discriminative ability, 1.0 indicated as having perfect discrimination, and values greater than 0.70 considered to reflect fair discrimination. The disease status of having hEDS was coded as a binary outcome variable, and the individual wearable metric values were treated as continuous predictors. Confidence intervals for ROC AUC were estimated using nonparametric bootstrap resampling. For each wearable metric, subjects were resampled with replacement 1000 times, and the AUC was recomputed for each resampled dataset. The 95% confidence interval was defined as the 2.5th and 97.5th percentiles of the bootstrap AUC distribution. (34)

### Principal Component Analysis

To explore the collective diagnostic potential of the physiological and survey-based metrics, an exploratory Principal Component Analysis (PCA) was performed. A consolidated data matrix was constructed using 19 variables, including: mean step count, nighttime means of LF, HF, LFHF ratio, PNN50, RMSSD, variability of LFHF ratio, SBP, DBP, step count, each PAGI-SYM domain score (Heartburn/regurgitation, Fullness/Early Satiety, Nausea/Vomiting, Bloating, Upper Abdominal Pain, Lower Abdominal Pain, and PAGI-SYM total), and lastly the Anxiety and Depression sub scores of the PROMIS-29. The input variables utilized different units of measurement; therefore, the data were z-score standardized to ensure variables with larger numerical ranges did not disproportionately influence the principal component weights. The PCA was executed using singular value decomposition of the standardized covariance matrix with the built in MATLAB function “pca()”. The contribution of each principal component (PC) was further evaluated with a scree plot.

### Statistics

Group comparisons of demographic variables were compared between hEDS and healthy cohorts using unpaired t-tests for continuous variables (age, BMI) and Chi-squared tests of independence for categorical variables (sex, ethnicity). For patient reported outcome measures (COMPASS-31, PAGI-SYM, PAGI-QOL, and PROMIS-29), group differences were evaluated using Mann-Whitney U tests. Wearable derived physiological metrics were compared between groups using independent samples t-tests. To evaluate the relationship between autonomic metrics and symptom burden, Spearman’s rank correlation coefficients with testing the hypothesis of no correlation against the alternative hypothesis of a nonzero correlation were calculated. For ROC analysis, to assess whether diagnostic performance was significantly better than chance, bootstrapping was employed to generate 95% confidence intervals (CIs), and significance was determined if the lower bound of the bootstrap CI exceeded 0.50. Statistical significance for all tests was defined as *p* < 0.05.

## RESULTS

### Participant Demographics

As shown in Table 1, the EDS and healthy control groups were not significantly different from each other in age (*p*-value: 0.97), BMI (*p*-value: 0.77), or sex. However, for race/ethnicity, there was a significant difference between cohorts (*p*-value: 0.01). Both groups consisted of all females. EDS, particularly hEDS, typically has a higher diagnosis frequency in females; however, it’s not clear whether a higher prevalence or rather difference in manifestation of symptoms in females contributes to this (35). Some subtypes of EDS do have a more balanced prevalence between males and females (35). However, this cohort specifically recruited individuals with an hEDS diagnosis, alongside age and sex matched controls.

**Table 1.**
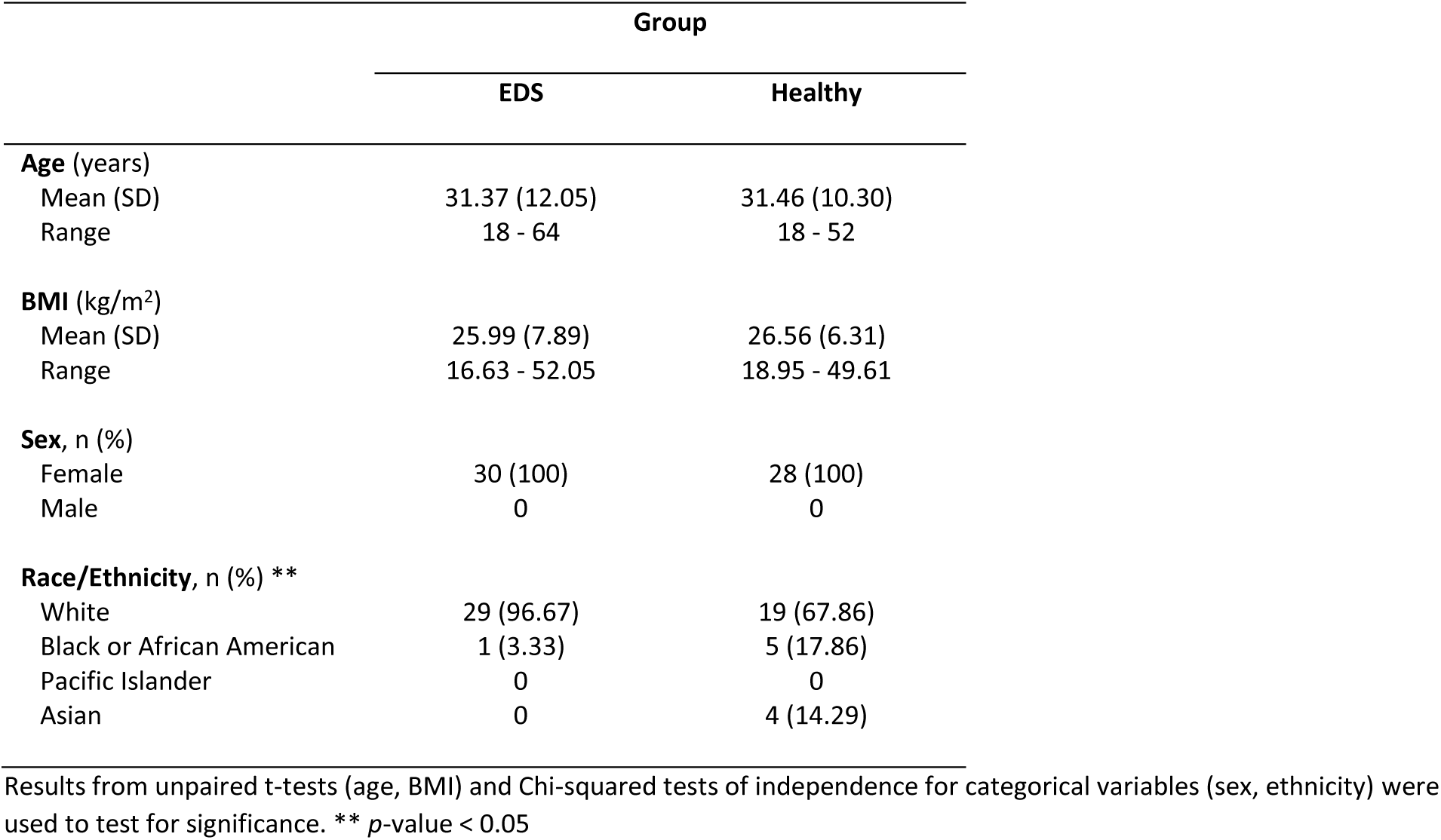
Study participant demographics.

### Symptom Surveys Results

Within the hEDS cohort, 11 individuals had a clinical diagnosis of gastroparesis prior, 19 did not.

However, based on their subjective symptoms at the time of completing the survey, a self-report of clinical classification of gastroparesis on the PAGI-SYM highlighted that 6 individuals reported “Vomiting-Predominant”, 24 reported “Dyspepsia-Predominant”, and 3 reported “Regurgitation-Predominant”; moreover, 2 individuals had reported both vomiting and dyspepsia, and 1 individual reported all three classifications. As summarized in Table 2, the hEDS cohort (n=30) demonstrated significantly greater symptom severity across all six PAGI-SYM domains compared to healthy controls (n=28; *p* < 0.001). The most pronounced symptoms in the hEDS group were fullness/early satiety (2.97 ± 1.26) and bloating (2.67 ± 1.53). Correspondingly, PAGI-QOL scores indicated that these gastrointestinal issues significantly impacted the quality of life for hEDS patients in all domains including daily activities, diet, psychological, clothing, and relationship (*p* < 0.001). In particular for hEDS cohort, diet and clothing had total QOL scores being markedly higher than those of the healthy cohort (2.23 ± 0.99 vs. 0.06 ± 0.16, 2.23 ± 1.60 vs. 0.11 ± 0.28).

**Table 2.** PAGI-SYM and PAGI-QOL Survey responses.

|  | PAGI-SYM |  |  |  | PAGI-QOL |  |  |
| --- | --- | --- | --- | --- | --- | --- | --- |
|  | hEDS<br>n=30 | Healthy<br>n=28 | *P*<br>Value |  | hEDS<br>n=30 | Healthy<br>n=28 | *P*<br>Value |
|  | Mean<br>(SD) | Mean<br>(SD) |  |  | Mean<br>(SD) | Mean<br>(SD) |  |
| Heartburn/regurgitation | 1.61(1.12) | 0.15(0.45) | <0.001 | DA-Daily activities | 2.13(0.99) | 0.06(0.16) | <0.001 |
| Fullness/early satiety | 2.97(1.26) | 0.14(0.26) | <0.001 | DI-Diet | 3.06(1.40) | 0.26(0.45) | <0.001 |
| Nausea/vomiting | 2.11(1.21) | 0.19(0.34) | <0.001 | PSY-Psychological | 1.75(1.15) | 0.16(0.43) | <0.001 |
| Bloating | 2.67(1.53) | 0.32(0.64) | <0.001 | CLO-Clothing | 2.33(1.60) | 0.11(0.28) | <0.001 |
| Upper abdominal pain | 2.42(1.03) | 0.09(0.24) | <0.001 | REL-Relationship | 1.21(1.00) | 0.00(0.00) | <0.001 |
| Lower abdominal pain | 2.18(1.13) | 0.04(0.19) | <0.001 |  |  |  |  |
| Total | 2.16(0.86) | 0.16(0.23) | <0.001 | Total | 2.23(0.96) | 0.13(0.24) | <0.001 |

P-values reflect results from Mann-Whitney U tests comparing distributions of survey responses between groups.

As shown in Table 3, the hEDS cohort (n=30) demonstrated significantly greater autonomic symptoms across all six COMPASS-31 domains (orthostatic, vasomotor, secretomotor, gastrointestinal, urinary, and pupillomotor) as well as total score compared to healthy controls (n=28; *p* < 0.001). Notably with hEDS cohort, the largest contrast to healthy controls were in the domains of orthostatic (25.20 ± 5.37 vs. 6.29 ± 8.53) and gastrointestinal (11.85 ± 4.22 vs. 2.49 ± 2.49).

**Table 3.** COMPASS-31 questionnaire scores.

|  | Group |  | *P*<br>Value |
| --- | --- | --- | --- |
|  | hEDS<br>n=30 | Healthy<br>n=28 |  |
|  | Mean (SD) | Mean (SD) |  |
| Orthostatic | 25.20(5.37) | 6.29(8.53) | <0.001 |
| Vasomotor | 2.33(1.66) | 0.21(0.77) | <0.001 |
| Secretomotor | 4.43(3.86) | 0.84(2.43) | <0.001 |
| Gastrointestinal | 11.85(4.22) | 2.49(2.49) | <0.001 |
| Urinary | 2.07(1.54) | 0.04(0.21) | <0.001 |
| Pupillomotor | 2.60(1.44) | 0.90(1.04) | <0.001 |
| Total | 48.48(12.34) | 10.77(11.70) | <0.001 |

P-values reflect results from Mann-Whitney U tests comparing distributions of survey responses between groups.

As shown in Table 4 from the PROMIS-29 survey, the hEDS cohort exhibited significantly impaired health status across all assessed domains compared to healthy controls. The most marked elevations in symptom burden were observed in fatigue (*p* < 0.001), pain interference (*p* < 0.001), and pain intensity (*p* < 0.001). Despite anxiety and depression score being close between the hEDS and healthy cohorts (57.37 ± 9.57 vs. 51.07 ± 9.08, and 53.58 ± 8.40 vs. 46.82 ± 7.28), they were both still significantly different between groups (anxiety: *p* = 0.016, depression: *p* = 0.003).

**Table 4.**
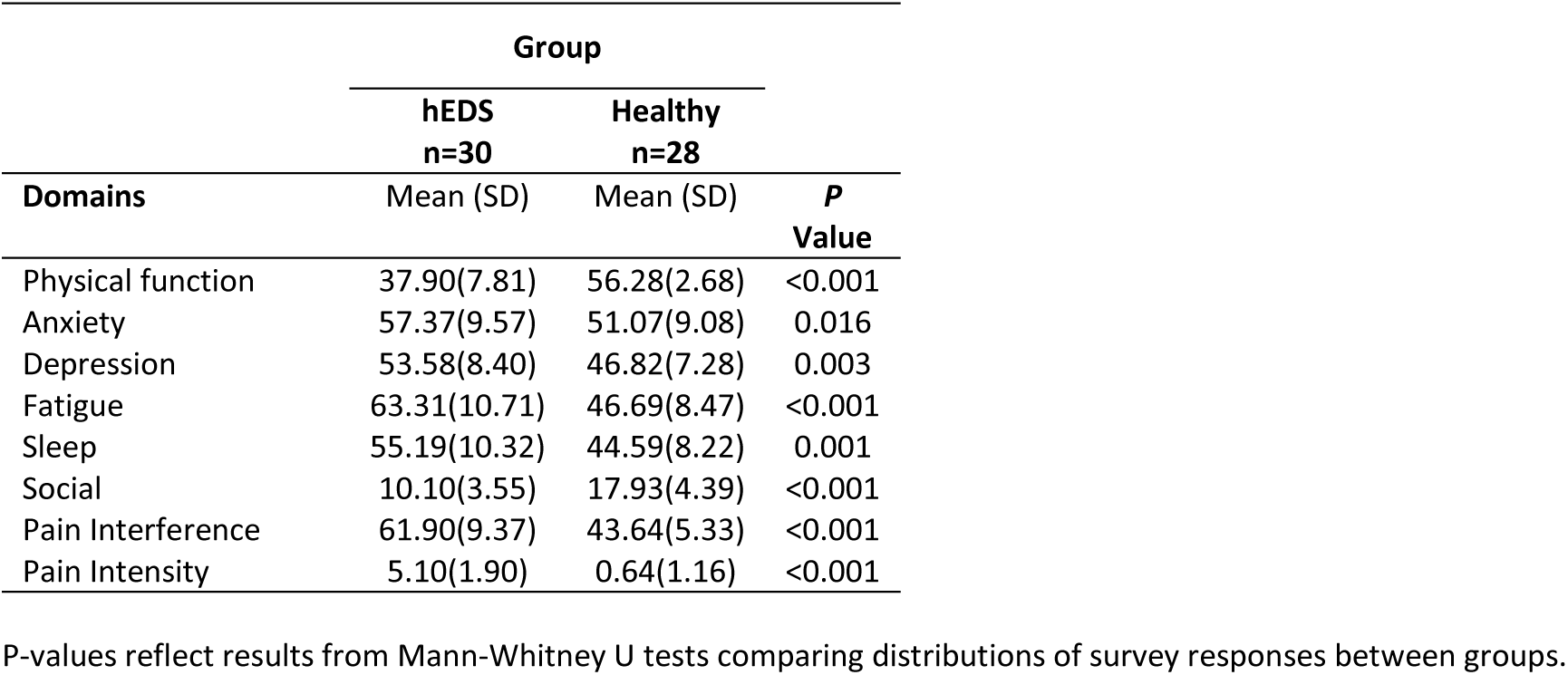
PROMIS-29 questionnaire scores.

### Wearable Metrics Results

To illustrate the utility of the long-term wearable monitoring, a physiological and symptomatic profile for a representative EDS participant is presented in Figure 1. Continuous 30-day sensor data (Panels A-D) demonstrated stable longitudinal trends in heart rate and blood pressure, while highlighting the significant degree of daily intra-individual variability. Aggregated circadian analysis (Panels B, D) further characterized these rhythms, revealing consistent nocturnal dipping patterns and diurnal fluctuations across the study duration. However, in this example individual, there was a characteristic inverse coupling between heart rate and RMSSD (Panel E), reflecting an atypical regulation of this parasympathetic HRV metric throughout the 24-hour cycle. Whereas with the example healthy participant in Figure 2, there is more an expected inverse nocturnal rise in HRV with concurrent fall in heart rate. These objective physiological patterns may be related with the participants’ self-reported clinical burden of COMPASS-31 and PAGI-SYM profiles (Panels F–G). Together, these two examples demonstrate feasibility of using multi-modal wearable platforms to capture the complex interplay between autonomic physiology and chronic symptom burden.

**Figure 1.**
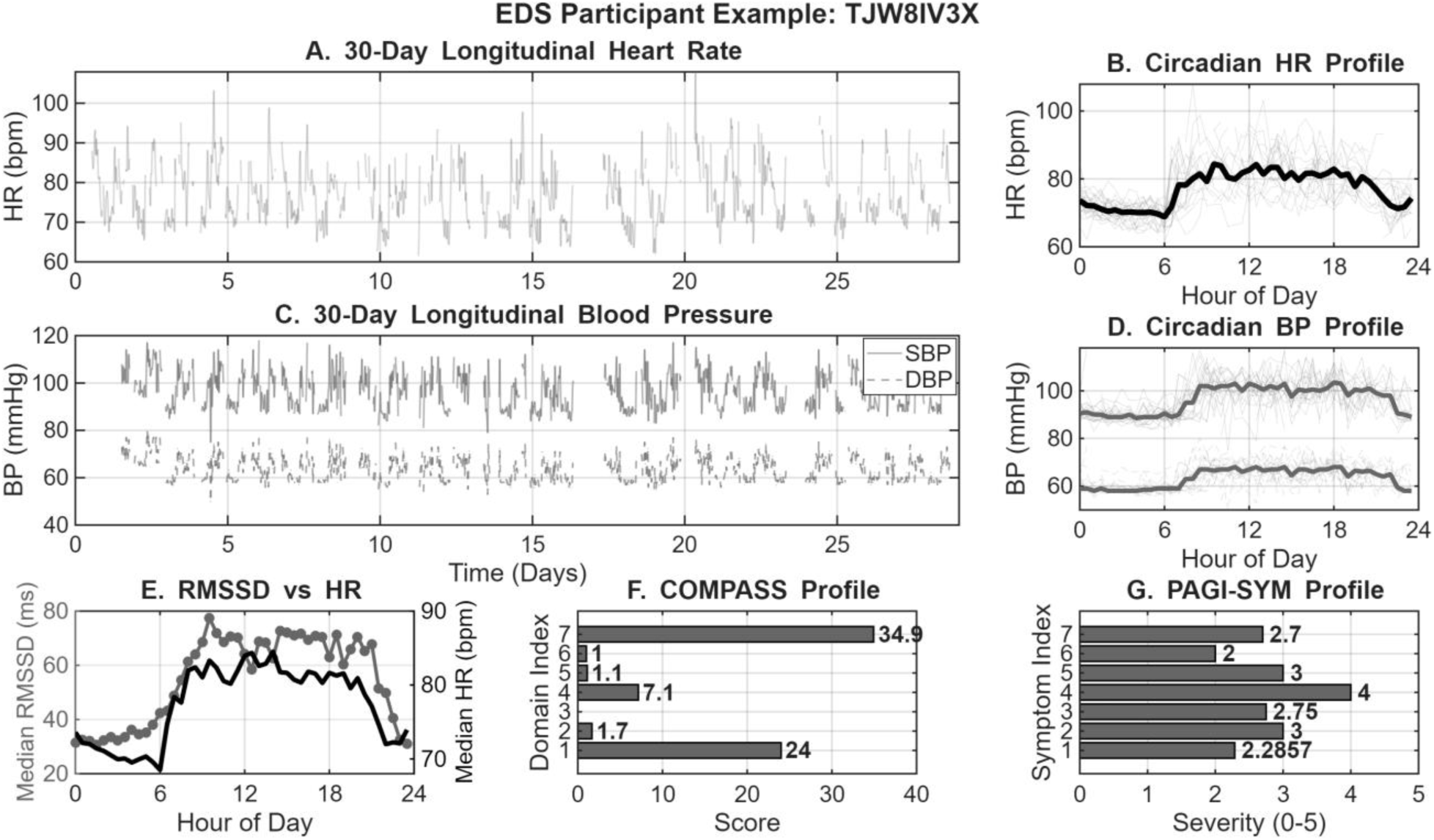
Integrated 30-day physiological and symptomatic profile for an EDS Participant. (A) Longitudinal heart rate (HR) displayed as 30-minute binned raw data. (B) Circadian HR profile showing overlapping daily cycles and the median diurnal rhythm (black). (C) Longitudinal systolic (SBP; solid) and diastolic (DBP; dashed) blood pressure (BP) trends over the study duration. (D) Median circadian BP profiles illustrating the diurnal variation of SBP and DBP. (E) Diurnal coupling between parasympathetic activity (median RMSSD; grey circles) and HR (black line). (F) Self-reported autonomic symptom burden across COMPASS-31 domains and (G) gastrointestinal symptom severity via the PAGI-SYM index (0–5 scale). In both survey plots, horizontal bar length indicates individual domain or symptom burden, where higher values represent greater clinical severity. If blank, then it indicates a score of 0.

**Figure 2.**
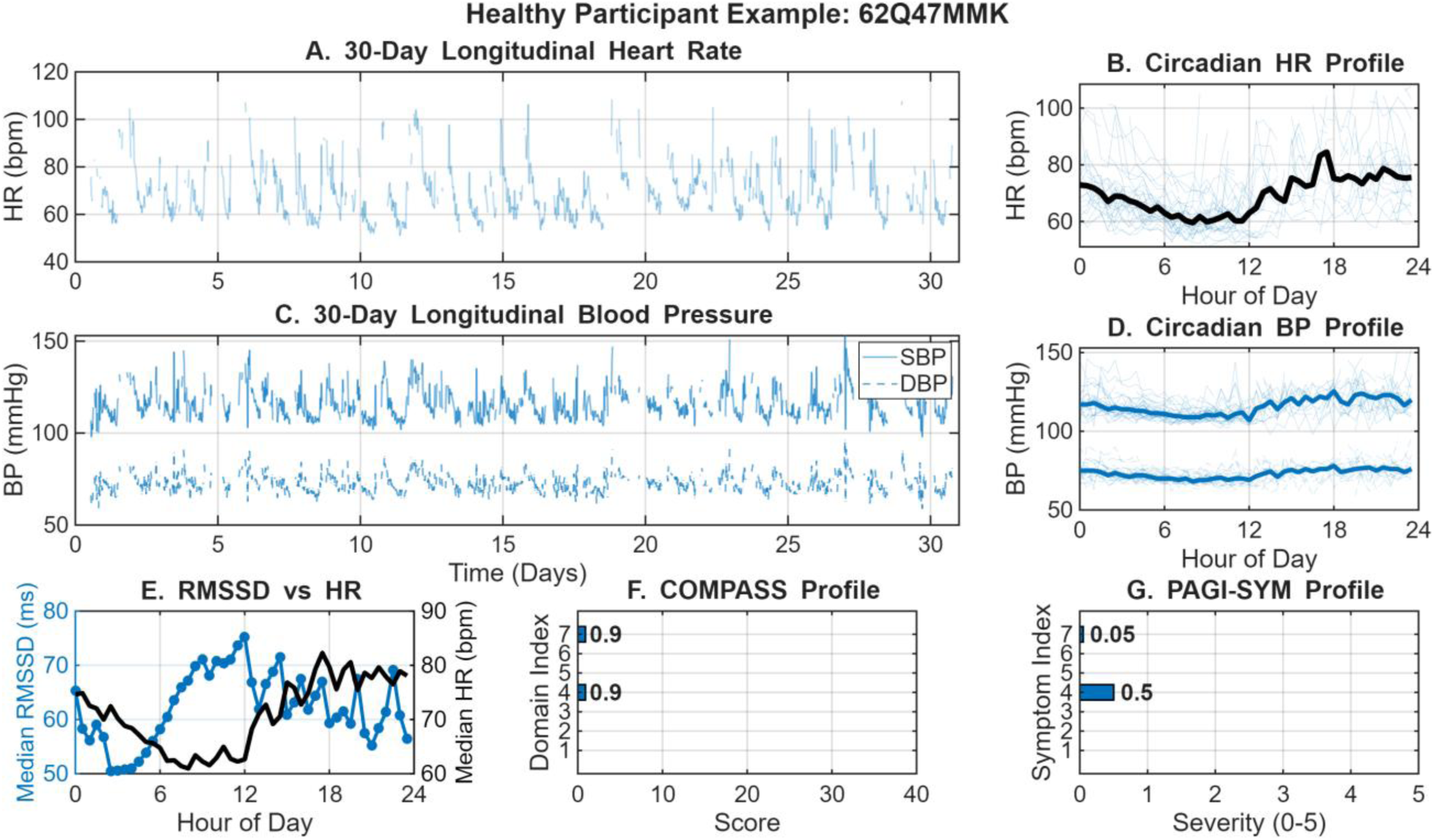
Integrated 30-day physiological and symptomatic profile for a Healthy Participant. (A) Longitudinal heart rate (HR) displayed as 30-minute binned raw data. (B) Circadian HR profile showing overlapping daily cycles and the median diurnal rhythm (black). (C) Longitudinal systolic (SBP; solid) and diastolic (DBP; dashed) blood pressure (BP) trends over the study duration. (D) Median circadian BP profiles illustrating the diurnal variation of SBP and DBP. (E) Diurnal coupling between parasympathetic activity (median RMSSD) and HR (black line). (F) Self-reported autonomic symptom burden across COMPASS-31 domains and (G) gastrointestinal symptom severity via the PAGI-SYM index (0–5 scale). In both survey plots, horizontal bar length indicates individual domain or symptom burden, where higher values represent greater clinical severity. If blank, then it indicates a score of 0.

There were little significant differences or trends when comparing means for the entire recording period (∼1 month) between the EDS and healthy cohorts. The only significant difference between groups was step count, where individuals with EDS have a significantly lower step count per 30 minute window (73.61 steps ± 33.22) compared to healthy controls (150.36 steps ± 96.18, *p* = 0.001).

However, when comparing means during nighttime rest between the hours 1:00–3:00 AM there were trends of note. As shown in Figure 3, LF and LF/HF ratio both trended higher at night in individuals with EDS as compared to healthy individuals. Furthermore, HF, PNN50, and RMSSD all trended lower at night in individuals with EDS as compared to healthy individuals. This suggests higher sympathetic activity and less parasympathetic activity during sleep in individuals with EDS compared to healthy controls.

**Figure 3.**
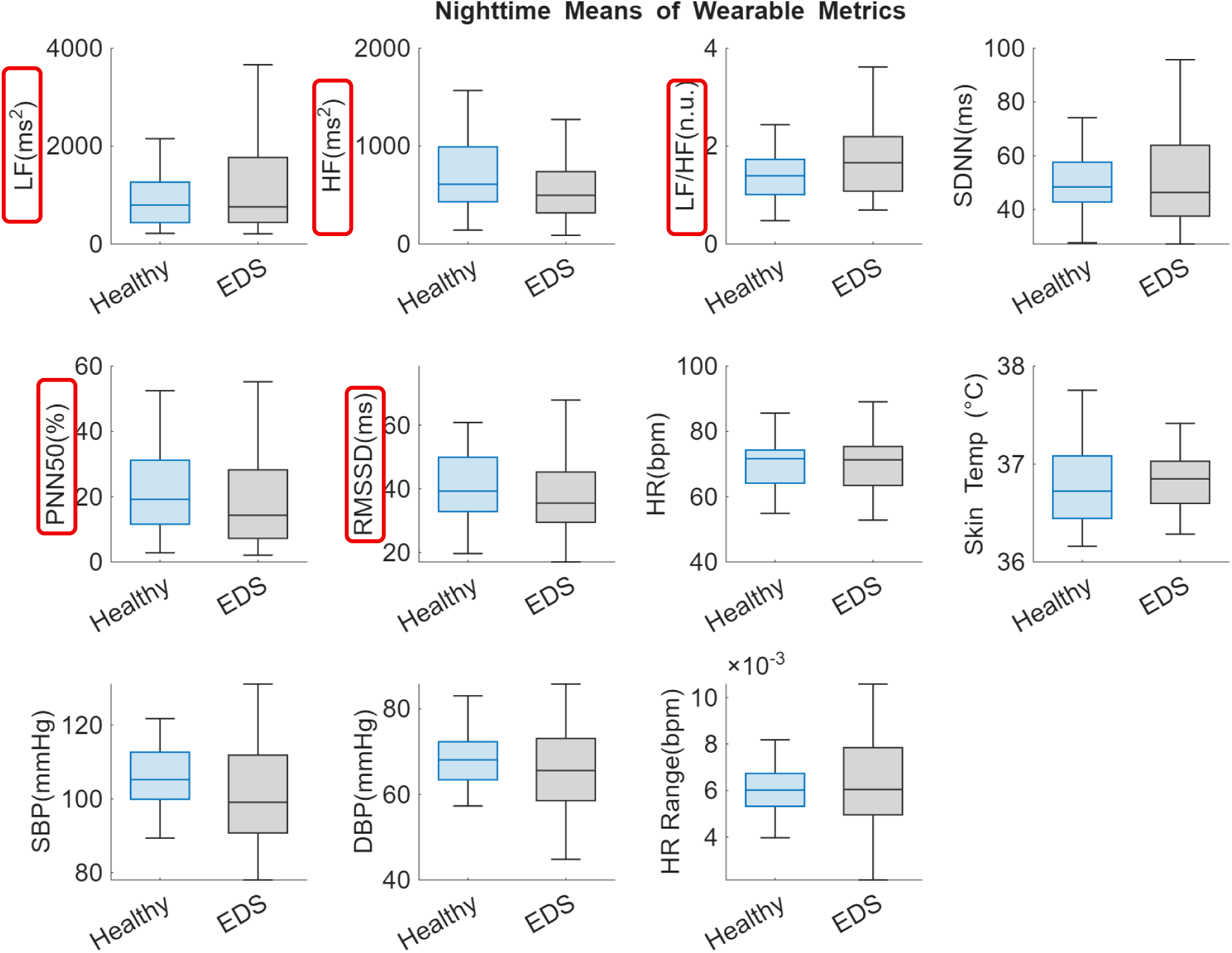
Comparison of nighttime wearable-based metrics between the healthy (blue) and EDS (grey) cohorts. Data were determined by averaging each metric from continuous recording during the hours of presumed nighttime (1:00–3:00 AM) over the entire recording period (∼30 days) for each individual. Bar plots show group means ± sem. There were no statistically significant differences (*p* < 0.05) observed compared with the two groups. However, LF and LF/HF ratio both trended higher at night in individuals with EDS as compared to healthy individuals. Additionally, HF, PNN50, and RMSSD all trended lower at night in individuals with EDS as compared to healthy individuals.

Additionally, when comparing stability of metrics during the same time windows across the ∼1 month recording period, there were significant differences between groups observed. As shown in Figure 4, there was much higher instability in the LF/HF ratio in the EDS cohort (SD= 0.68 ± 0.18) compared to the healthy cohort (SD = 0.58 ± 0.18, *p* = 0.044). Both systolic and diastolic blood pressure had higher instability in the EDS cohort (SD= 7.60 ± 3.17, 5.14 ± 1.90) compared to the healthy cohort (SD = 6.33 ± 0.66, 4.30 ± 0.38, *p* = 0.043 & 0.026). Lastly, the step count had much less variation, more stability in the EDS cohort (SD= 100.59 ± 40.83) as compared to the healthy cohort (SD = 238.02 ± 146.15, *p* < 0.001). When examining further into hours of this stability, as shown in Figure 5, most significant differences in LF/HF ratio variability occur during the night, approximately between 2:00–10:00 AM. Whereas most of the SBP significant variability difference between groups occurs during wake hours of approximately 12:00–8:00 PM. These findings suggest that blood pressure and sympathovagal balance are much less stable in the EDS group compared with healthy controls.

**Figure 4.**
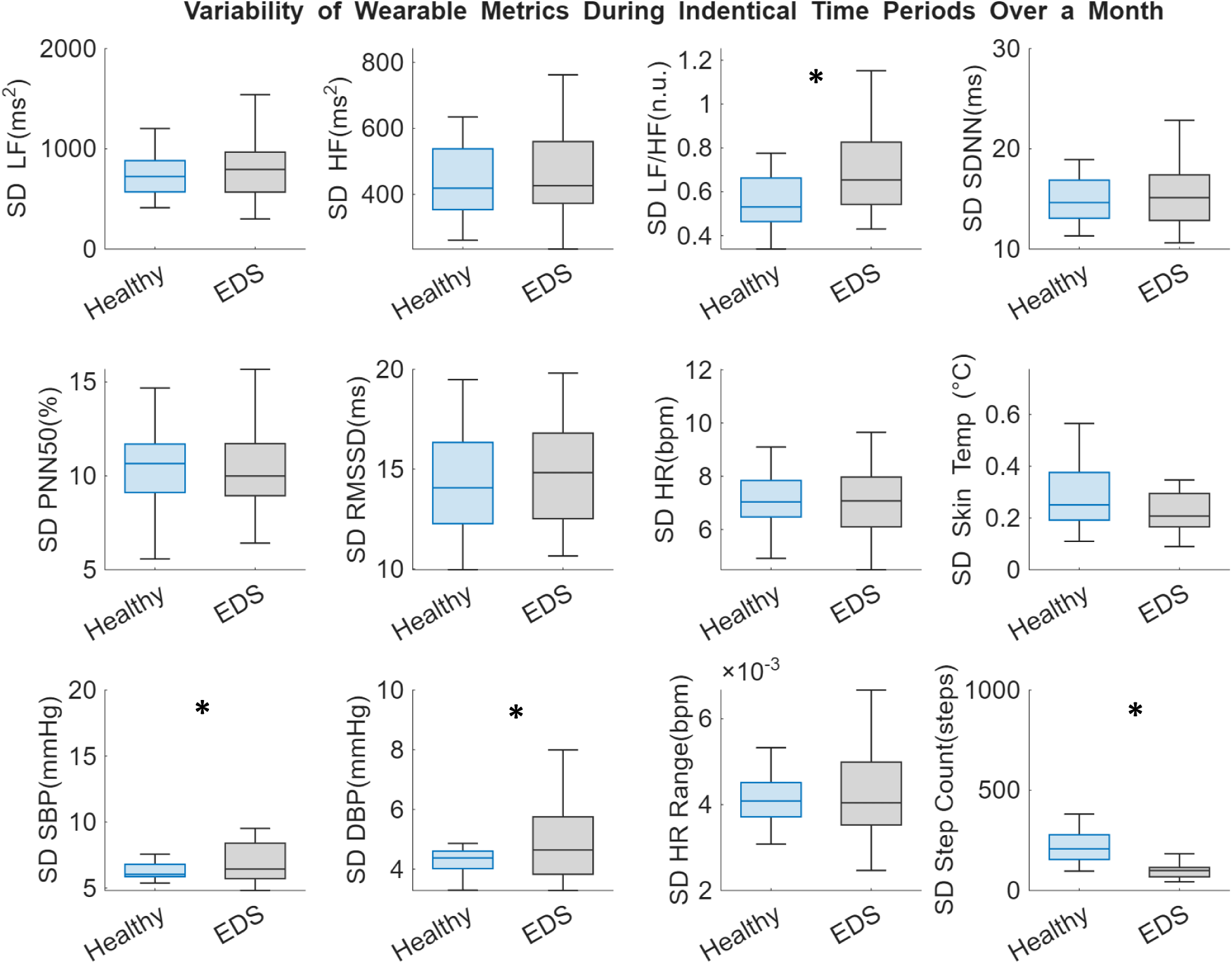
Comparison of wearable-based metrics between the healthy (blue) and EDS (grey) cohorts. Data were determined by binning data for each metric into 30 minute time windows on a 24 hour scale over the entire recording period (∼30 days) for each individual. Then, for each time window, the standard deviation was calculated to represent variability within that time window across the month. Bar plots show group means ± sem. There were statistically significant differences (*p* < 0.05) observed in LF/HF ratio (*p* = 0.044), SBP (*p* = 0.043), DBP (*p* = 0.026), and step count (*p* < 0.001).

**Figure 5.**
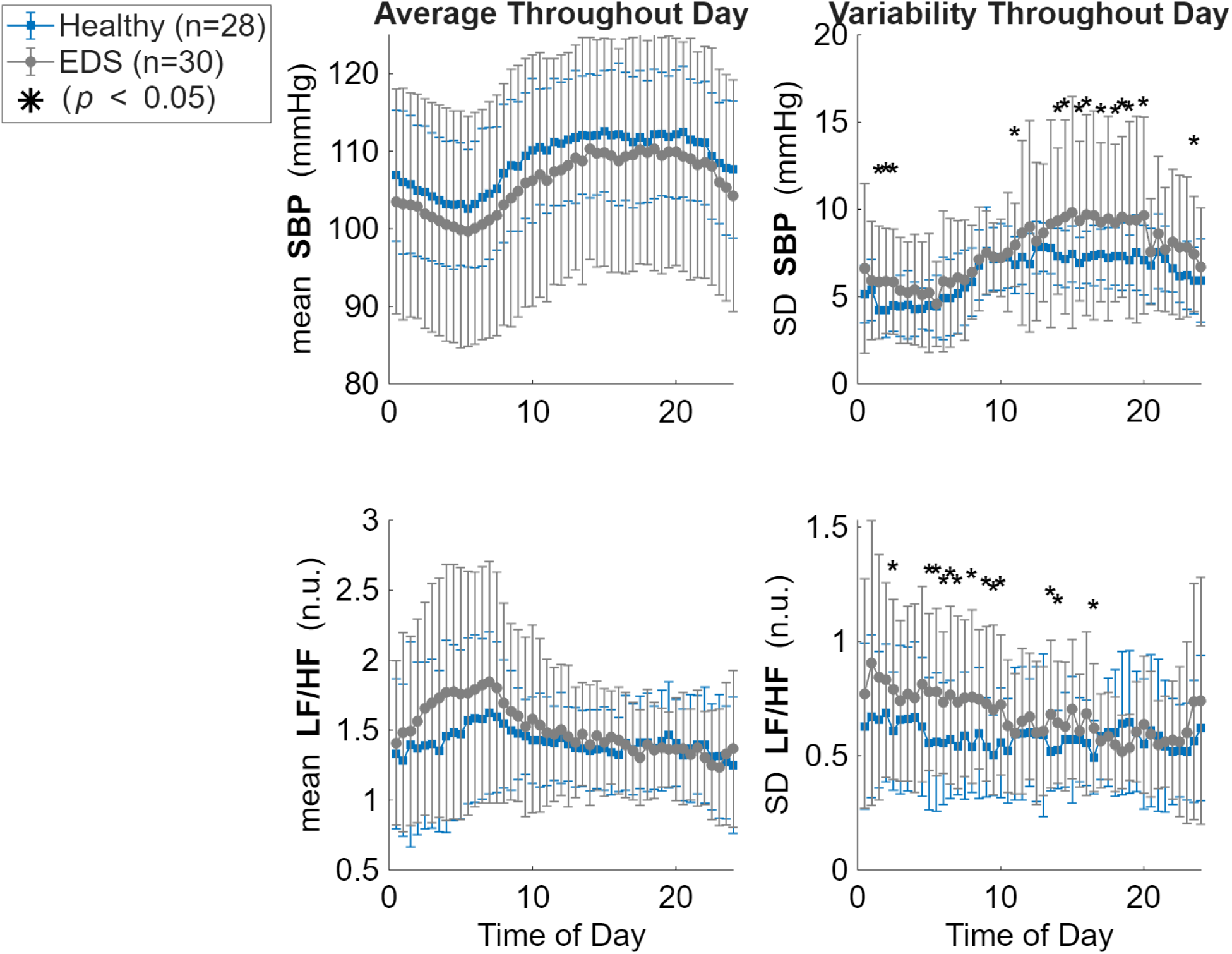
Comparison of wearable-based metrics between the healthy (blue) and EDS (grey) cohorts. Data were determined by binning data for each metric into 30-minute time windows on a 24-hour scale over the entire recording period (∼30 days) for each individual. Then, for each time window, the average and standard deviation was calculated. Each point show group means ± SD for each time interval. There were statistically significant differences (*p* < 0.05) observed in variability of LF/HF ratio and SBP, but not in their means.

### Correlation Results

In the healthy cohort, there were many significant correlations between wearable physiological measurements and their reported survey outcomes as shown in Figure 6. Of note, primarily, mental health related outcomes were negatively correlated with autonomic metrics such as LF (rho =-0.47), SDNN (rho =-0.42), and SBP (rho =-0.40) significantly negatively correlated with the psychological sub score of PAGI-SYM. As well as LF (rho =-0.40) and SDNN (rho =-0.41) significantly negatively correlated with PAGI-QOL total score. Additionally, RMSSD (rho = 0.41) and PNN50 (rho = 0.39) were significantly positively correlated with PROMIS-31’s physical function sub score and anxiety sub score respectively. Furthermore, while not significant, higher correlation values were seen with autonomic metrics and anxiety and depression sub scores. In contrast, for the EDS cohort, significant correlations more so existed between autonomic metrics and PAGI-SYM reported outcomes, as opposed to more mental health focused outcomes as seen with the healthy controls. As shown in Figure 7, lower abdominal pain was significantly correlated with LF (rho = 0.52), HF (rho = 0.60), SDNN (rho = 0.51), RMSSD (rho = 0.44), and HR (rho = 0.38). Additionally, bloating was significantly correlated with LF (rho = 0.37), and fullness/satiety correlated with HR (rho = 0.50). Furthermore, HR correlated with nausea/vomiting (rho = 0.57) as well was PAGI-SYM total score (rho = 0.38). Overall, in the EDS cohort there appears to be a relationship between GI related reported outcomes and their wearable physiology, were as with the healthy cohort the relationship is between mental health reported outcomes and their wearable physiology.

**Figure 6.**
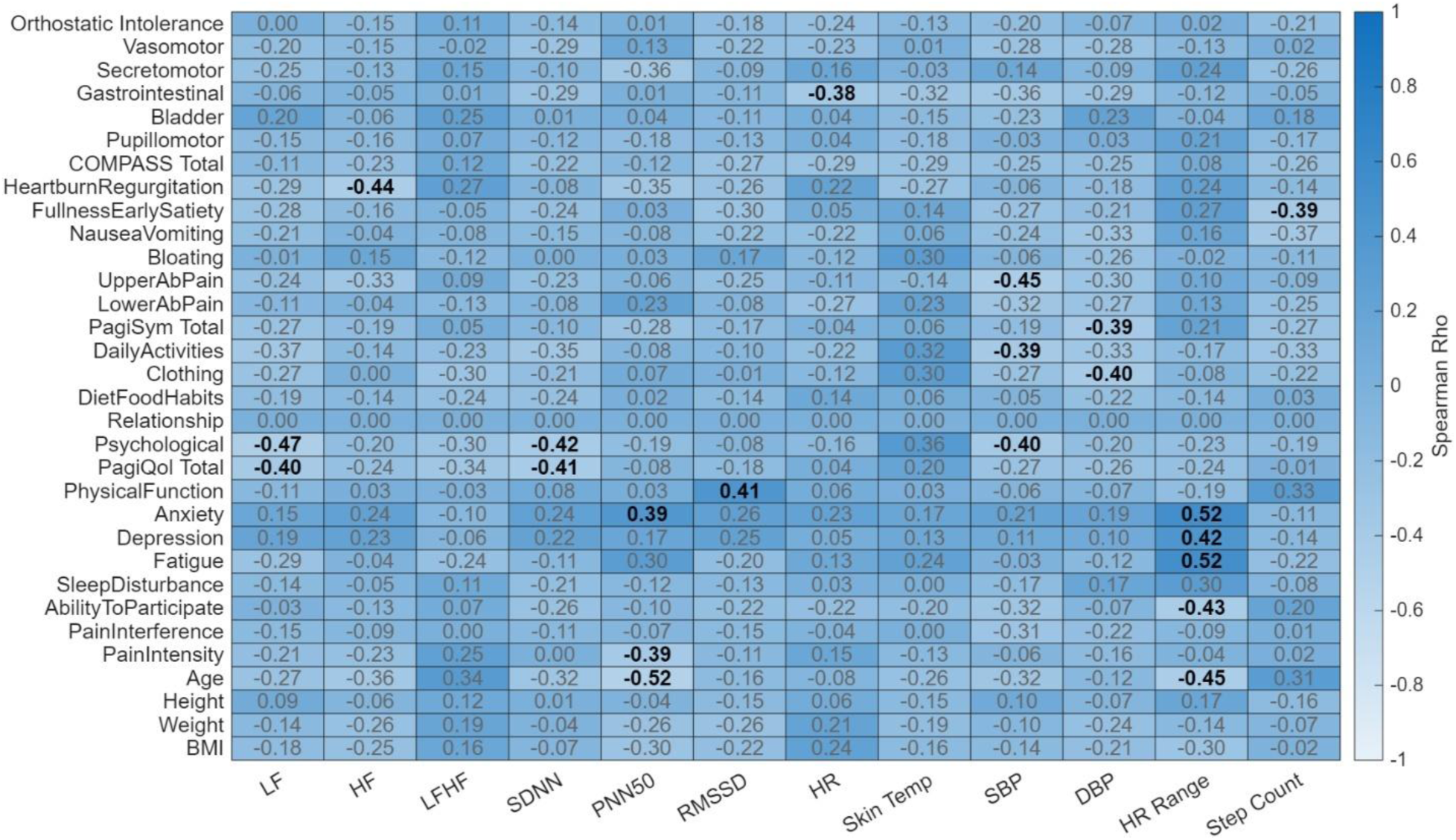
Heatmap of Spearman rank correlations between wearable physiological metrics and clinical/demographic profiles in the healthy cohort. The heatmap illustrates the relationship (rho) between wearable derived metrics and participant characteristics. Column variables encompass physiological wearable markers, including heart rate variability (LF, HF, LF/HF, SDNN, pNN50, RMSSD), heart rate (HR, HR Range), skin temperature, blood pressure (SBP, DBP), and step count. Row variables consist of demographic data (age, height, weight, and BMI) and comprehensive clinical assessments, including sub scores and total scores from the Composite Autonomic Symptom Score 31 (COMPASS-31), Patient Assessment of Upper Gastrointestinal Disorders-Symptom Severity Index (PAGI-SYM), Patient Assessment of Upper Gastrointestinal Disorders-Quality of Life (PAGI-QOL), and Patient-Reported Outcomes Measurement Information System (PROMIS-29). The color gradient represents the strength and direction of the correlation, with the color bar indicating Spearman’s Rho values ranging from-1.0 to 1.0. Correlation coefficients are overlaid on each cell, where bold text denotes statistical significance (*p* < 0.05) and gray text indicates non-significant associations.

**Figure 7.**
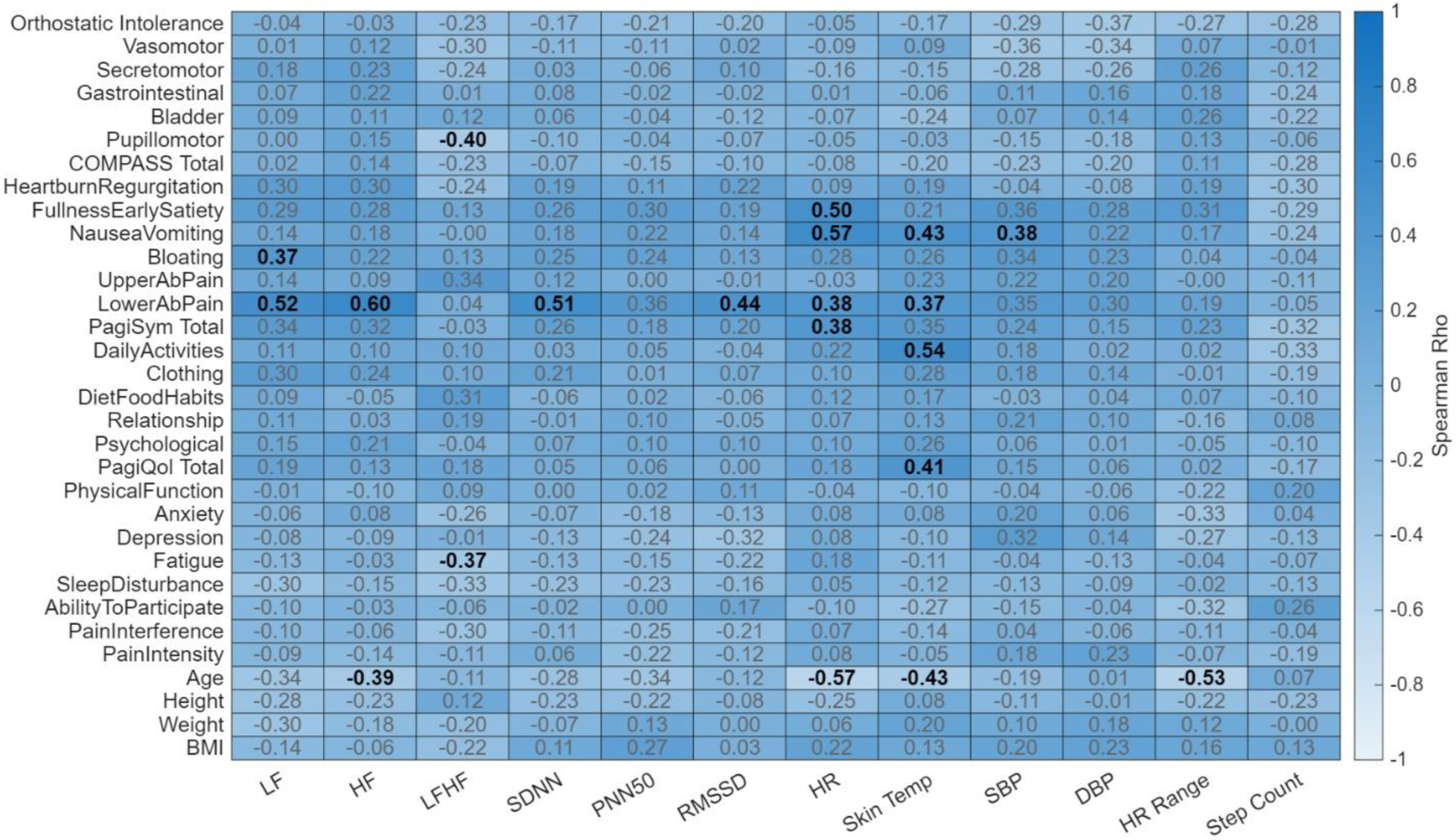
Heatmap of Spearman rank correlations between wearable physiological metrics and clinical/demographic profiles in the EDS cohort. The heatmap illustrates the relationship (rho) between wearable derived metrics and participant characteristics. Column variables encompass physiological wearable markers, including heart rate variability (LF, HF, LF/HF, SDNN, pNN50, RMSSD), heart rate (HR, HR Range), skin temperature, blood pressure (SBP, DBP), and step count. Row variables consist of demographic data (age, height, weight, and BMI) and comprehensive clinical assessments, including sub scores and total scores from the Composite Autonomic Symptom Score 31 (COMPASS-31), Patient Assessment of Upper Gastrointestinal Disorders-Symptom Severity Index (PAGI-SYM), Patient Assessment of Upper Gastrointestinal Disorders-Quality of Life (PAGI-QOL), and Patient-Reported Outcomes Measurement Information System (PROMIS-29). The color gradient represents the strength and direction of the correlation, with the color bar indicating Spearman’s Rho values ranging from-1.0 to 1.0. Correlation coefficients are overlaid on each cell, where bold text denotes statistical significance (*p* < 0.05) and gray text indicates non-significant associations.

### ROC Results

To evaluate the potential diagnostic utility of the wearable-derived parameters, ROC curve analysis was performed, as shown in Table 6. The most robust predictive power was observed for LFHF variability and SBP nighttime variability, which yielded AUC values of 0.68 (95% CI: 0.53-0.81) and 0.68 (95% CI: 0.53–0.82), respectively. The six top performing physiological metrics demonstrating the greatest ability to discriminate between hEDS and healthy cohorts, are visualized in Figure 8. The LFHF variability and SBP nighttime variability were the only AUC values that were considered statistically significant since the lower bound of the 95% bootstrap confidence interval exceeded 0.5, indicating performance greater than chance. Overall, these results suggest that no sole wearable metric alone has excellent capability to distinguish individuals with hEDS from healthy controls.

**Figure 8.**
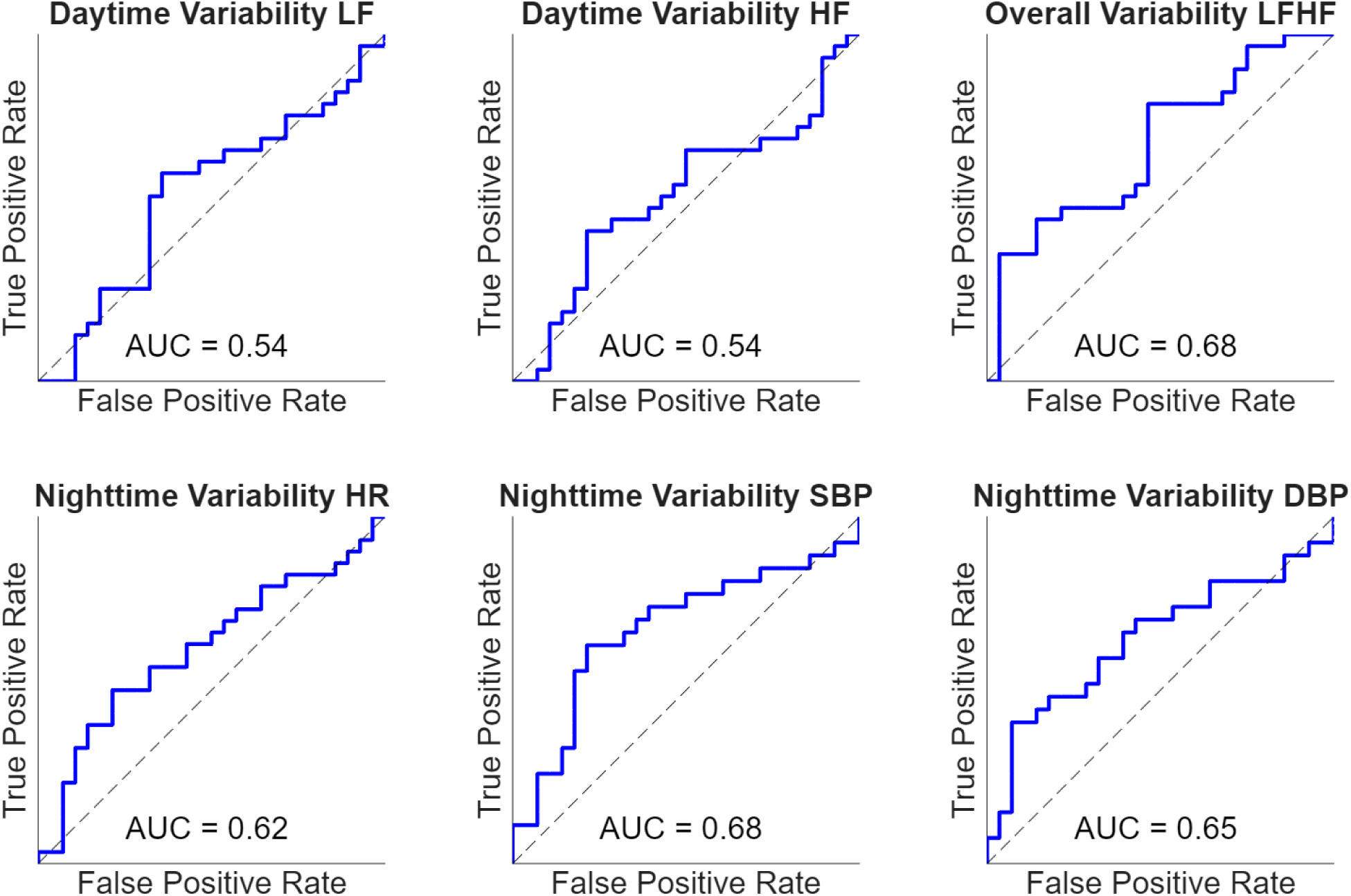
Receiver operating characteristic (ROC) curves assessing the discriminative ability of individual wearable derived day-night values to distinguish between participants with hEDS and healthy controls. Area under the curve (AUC) values are presented for each metric. These analyses were exploratory and intended to assess feasibility for future studies.

**Table 6.**
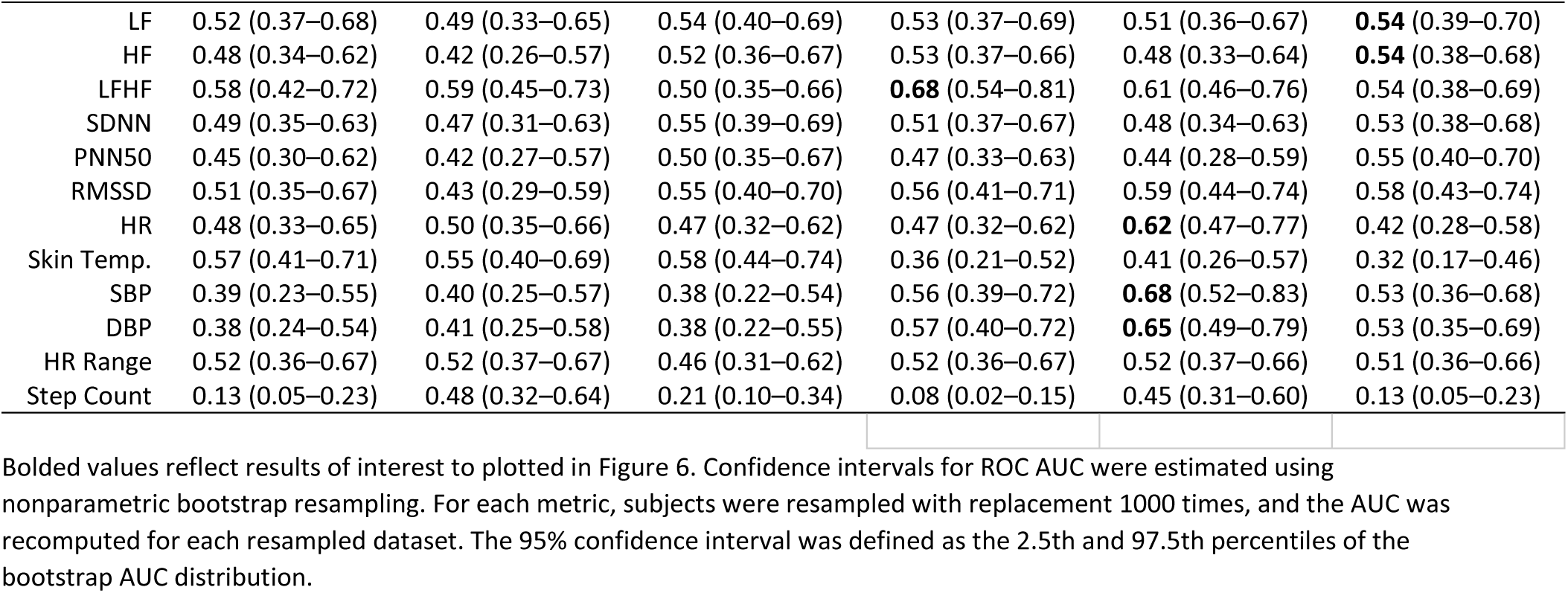
ROC Area Under Curve (AUC) values with 95% CI.

### PCA Results

To further explore the diagnostic potential of the wearable and self-reported symptom outcome profiles, an exploratory Principal Component Analysis (PCA) was conducted on 19 assessed parameters of interest based upon the prior analyses above. These parameters were: mean step count, nighttime means of LF, HF, LFHF ratio, PNN50, RMSSD, variability of LFHF ratio, SBP, DBP, step count, each PAGI-SYM domain score (Heartburn/regurgitation, Fullness/Early Satiety, Nausea/Vomiting, Bloating, Upper Abdominal Pain, Lower Abdominal Pain, and PAGI-SYM total), and lastly the Anxiety and Depression sub scores of the PROMIS-29. These specific parameters were selected based upon wearable derived trends discussed above as well as the unique correlations present in the cohorts between autonomic wearable metrics and reported stressors of either GI related symptoms or mental health related symptoms. The analysis revealed that the majority of the total variance was captured by a small number of components, with the first six principal components (PCs) accounting for 90.4% of the cumulative variance as shown in Figure 10. No clustering techniques were performed for this analysis as it was pure exploratory and a pilot sample size. However, there is clear separation visualized by these initial PCs, shown in Figure 9, suggesting that the multidimensional profile in hEDS offer strong potential for diagnostic classification or screening.

**Figure 9.**
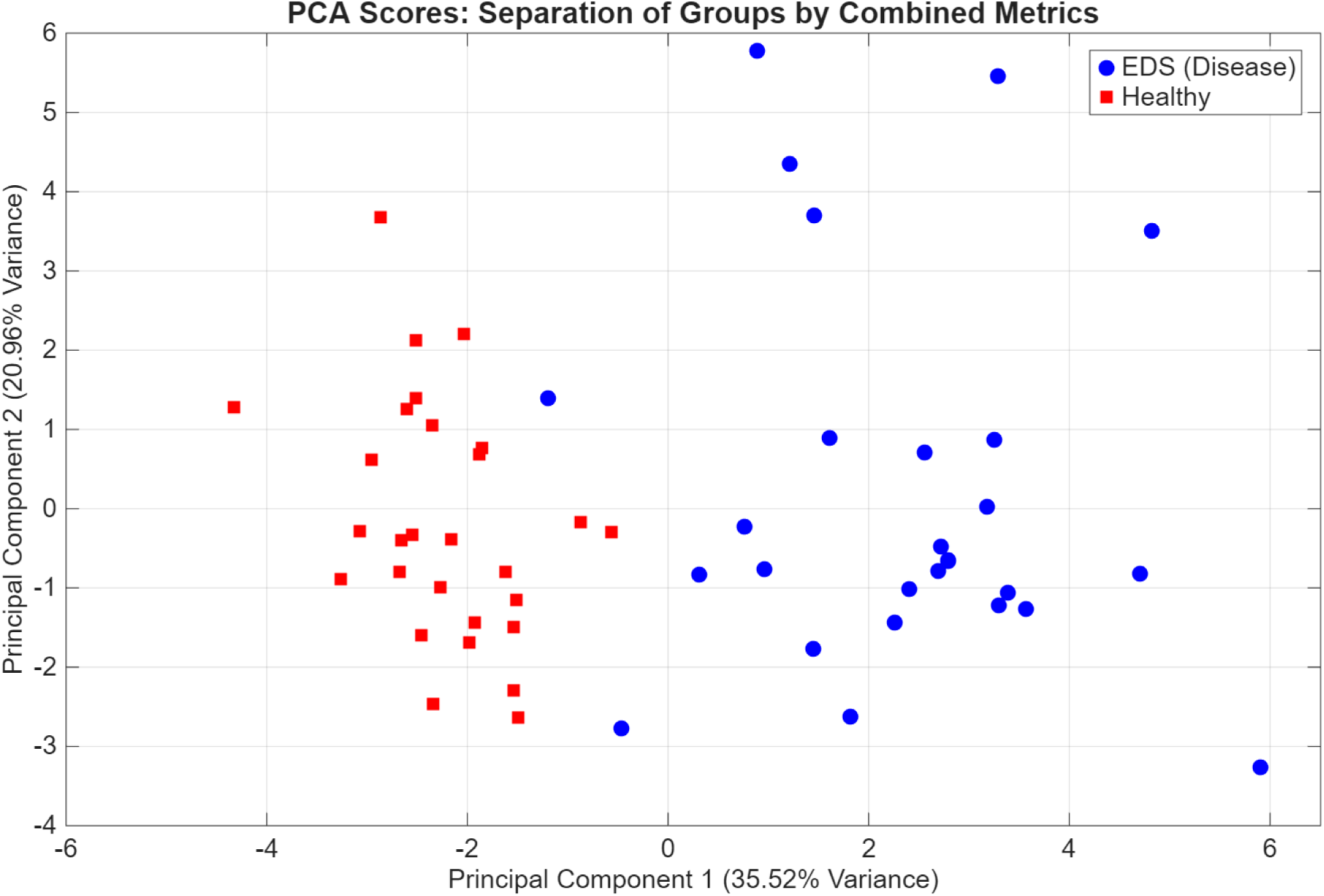
PCA plot of the first two principal components. Individual participants are plotted according to their scores for PC1 and PC2, which explain 35.52% and 20.96% of the total variance, respectively. Healthy controls (red squares) and individuals with hEDS (blue circles) demonstrate distinct spatial clustering, indicating a clear separation in multidimensional profiles between the two cohorts.

**Figure 10.**
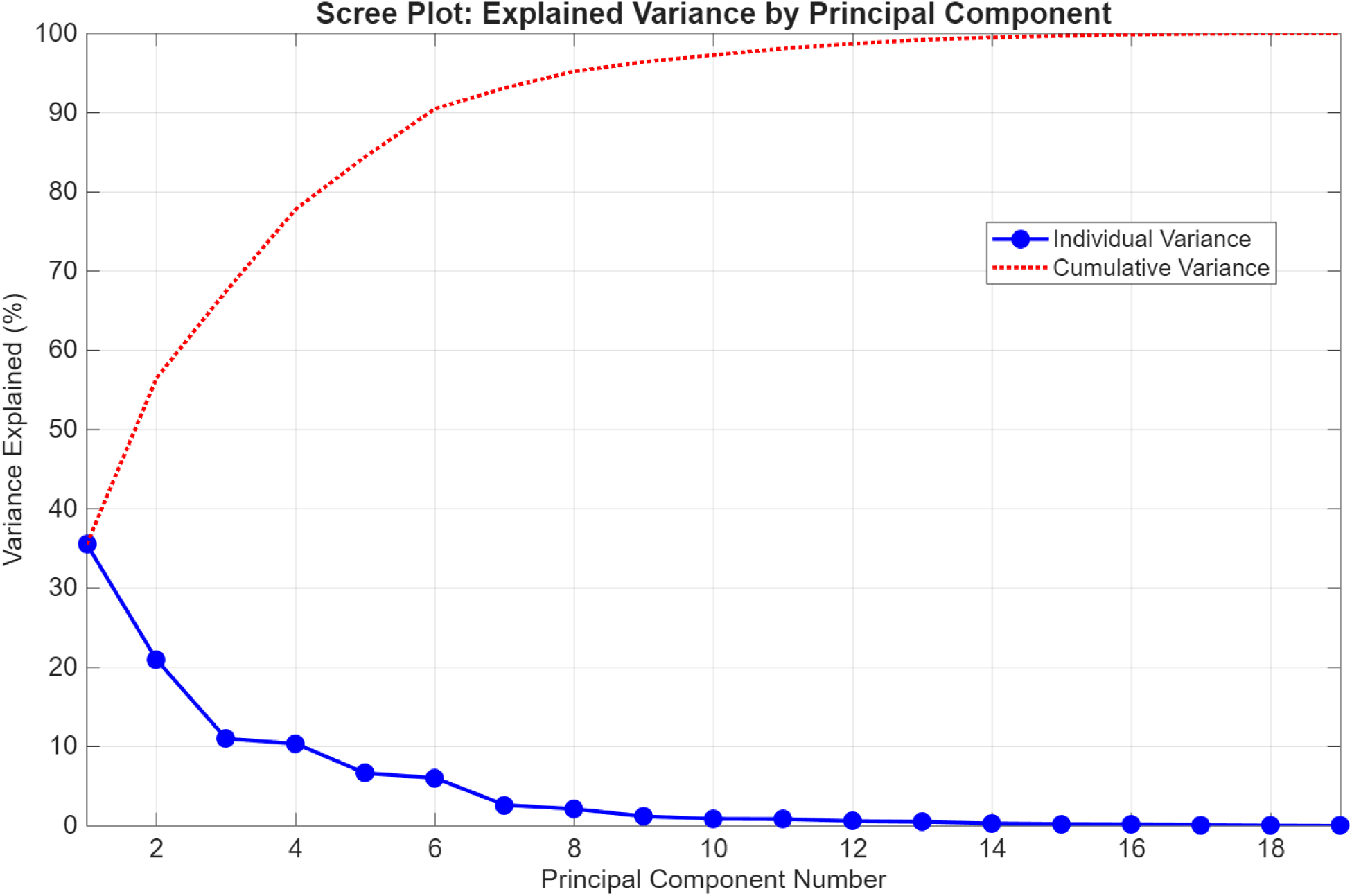
Scree plot of principal components and cumulative variance. The first six components reached 90.4% of the total variance explained across the 19 assessed parameters.

## DISCUSSION

The objective of this study is to evaluate circadian autonomic function from wearables and investigate their relationship with symptoms and to explore the feasibility of using wearable derived metrics to distinguish between individuals with and without hEDS. Overall, differences were discovered in wearable derived autonomic function and a unique relationship between reported symptoms and wearable physiology was identified for both groups, healthy and hEDS.

### Wearable-based Metrics

Manifestations of hEDS are highly variable and can range from one organ system dominating symptoms to multiple systems (9,11). Furthermore, country of residence has also been described to influence symptomology (6).Therefore, it’s important to consider this is a single site study in the United States, which may affect wide applicability of the findings in the paper. Furthermore, due to the wide variance in manifestations, it was not unexpected that significant differences from wearable tracking may not be strong given the sample size of roughly 30 participants per cohort. Given that physical disability is high in hEDS, it was expected that step counts would be significantly lower in the hEDS cohort as compared to the healthy cohort (11). Furthermore, when observing the variability or stableness of the step counts throughout the study period of ∼30 days, it also was expected that step counts would not vary much in the hEDS cohort compared to healthy cohort.

While not significant, the trends when observing nighttime metrics suggest a sympathetic dominance or parasympathetic withdrawal with LF, LF/HF ratio both trended higher, and HF, PNN50, and RMSSD trended lower at night in EDS. Additionally, HF, PNN50, and RMSSD all trended lower. These findings are consistent with De Wandele et al. who found hEDS participants showed increased sympathetic activity at rest when undergoing autonomic testing in a clinical setting (13). When viewing the stability or variability by time window of the autonomic metrics, our results showed blood pressure and sympathovagal balance are much less stable in the hEDS group compared with healthy controls.

This is expected as it is well known there is autonomic, specifically cardiovascular, dysfunction prevalent in hEDS (12,36). This work is among the first to utilize a wrist-based wearable for ambulatory blood pressure tracking, specifically in this autonomic dysfunction population. The instability of the systolic and diastolic values, particularly in the hEDS cohort highlight a unique but expected observation within hEDS given the previously studied baroreflex sensitivity compared to controls (13,36).

To our knowledge this is the first published experiment comparing wearable-based data in hEDS compared to healthy matched controls. While Mathena et. al utilized wearable tracking in hEDS, there were no matched controls to compare wearable metrics on a group level (21). However, Mathena et. al did observe a relationship between the HRV parameters VLF and ULF with days where GI-symptoms were reported high (21). Our study did not have rigorous daily symptom reporting nor the same length of study period (∼120 days) as their study; therefore, we are unable to directly relate our results.

However, like them, we found a significant relationship between autonomic wearable metrics and GI reported symptoms.

In general, insights into autonomic physiology captured with wearable technology offer significant potential as supplements or alternatives to traditional autonomic diagnostics. Standard clinical assessments, such as office based orthostatic heart rate and blood pressure, head-up tilt table testing (HUTT) in cardiology, and formal autonomic reflex testing in neurology, frequently yield false negatives in patients with hypermobile Ehlers-Danlos Syndrome (hEDS) (37). This diagnostic gap often stems from the highly controlled nature of clinical settings, which provide only a brief’snapshot’ of cardiovascular behavior. Consequently, these one-time assessments often fail to capture the transient yet debilitating autonomic dysfunction that characterizes the daily lived experience of individuals with hEDS. Wearables hold immense potential to be used in combination or alone in closing the gap seen in many conditions, like hEDS, for monitoring, screening, and diagnosis (38).

### Survey Outcomes

Every reported symptom survey score and sub-score were significantly different between healthy and hEDS cohorts. This finding was expected given the debilitating multifaceted nature of the hEDS condition and quality of life. Our findings for PROMIS-29 anxiety and depression match very well with Bieniak et. al’s findings in their pediatric hEDS cohort (39). Furthermore, all scores of PROMIS-29 also aligned with Estrella and Frazier’s findings, other than the social and pain interference scores, which greatly differed from our findings (40). Our COMPASS-31 scores for hEDS were similar to prior work by Hertel et. al done with 70 individuals with hEDS (41). PAGI-SYM reported findings aligned with prior work observing patients with dyspepsia and gastroparesis, both of which are frequent among hEDS patients (42,43). To our knowledge, these are the first published reporting of PAGI-QOL results in hEDS. PAGI-QOL scores in the hEDS cohort were expected to be significantly different than the healthy control cohort due to the widely documented GI complications with hEDS.

A compelling finding in this study was the contrast in correlations between autonomic wearable-derived metrics and patient reported symptoms across the two cohorts. In the healthy cohort, changes in these autonomic metrics were primarily associated with psychological indices, including the PROMIS-29 anxiety and depression subscales and the PAGI-QOL psychological domain and total score. This correlation aligns with the heavily studied influence of mental health burden and generalized stress on autonomic nervous system modulation in general. (44–47)

In contrast, the autonomic wearable metrics in the hEDS cohort were uniquely uncorrelated or decoupled from psychological symptom reports and were instead strongly correlated with gastrointestinal (GI) specific symptoms as measured by the PAGI-SYM. While overall survey scores for anxiety and depression were significantly higher in the hEDS group, these metrics did not appear to correlate or drive the autonomic response. This suggests that in hEDS, the primary stressor to the ANS may be a physiological burden, specifically GI related as opposed to a psychological state. As a consequence, the elevated psychological distress observed in hEDS may represent a secondary response to chronic symptom burden rather than the principal driver of autonomic changes measured with the wearable. It is also possible that the GI symptoms drive a stress state response and are what is causing the autonomic wearable watch findings, and possibly not hEDS specifically itself. It is important to state there needs to be future comparison with cohorts of both hEDS without GI symptoms and with normal gastric emptying to further understand and elucidate this relationship. Furthermore, a comparison between individuals with hEDS and individuals with hypermobility spectrum disorder who do not meet criteria of hEDS would further help add to our understanding of discriminatory power in these wearable findings and correlations.

### Diagnostic and Screening Potential

ROC results showed few metrics that had any diagnostic distinguishing capability on their own. This was expected due to the vast symptom presentation and diversity within the hEDS population, as well as our size of cohort. LFHF ratio stability was one of the few that showed it had meaningful discrimination. To our knowledge, there has been one other study exploring this idea of stability or fluctuations in the LFHF ratio with a wearable. Kishimoto et. al found that the LFHF ratio fluctuations were not different when comparing workers based on reported low or high well-being (48). While not a simple comparison, it may support our hypothesis of generalized environmental stress affecting sympathovagal balance and autonomic metrics differently from GI related stress, as we found the hEDS cohorts’ fluctuations, or instability, in LFHF ratio to be much higher than the healthy cohort. However, it is important to consider that LFHF ratio may not directly represent sympathovagal balance, as that line of thought has been under debate and its usefulness as a metric has been challenged (29,49). SBP and DBP stability during nighttime showed some diagnostic distinguishing capabilities. While measuring non-invasive blood pressure from a wearable in an extended time setting is a new emerging technology, we think our findings align with what has been found with hEDS prior. Since impaired blood pressure and heart rate control are very common in hEDS, it logically follows that blood pressure fluctuations would be significantly higher in an hEDS cohort compared to a healthy cohort, especially during the night where wearable readings are most accurate (35,36,41). However, it is also important to consider these non-invasive blood pressure values, while measured with a clinically validated method, are still only every 30 minutes and thus may not capture rapid, transient changes.

Leveraging these comparisons in wearable analyses and survey score correlations, a combinatory screening diagnostic approach was attempted with our Principal Component Analysis (PCA). Our PCA analysis leveraged 19 metrics (mean step count, nighttime means of LF, HF, LFHF ratio, PNN50, RMSSD, variability of LFHF ratio, SBP, DBP, step count, each PAGI-SYM domain score (Heartburn/regurgitation, Fullness/Early Satiety, Nausea/Vomiting, Bloating, Upper Abdominal Pain, Lower Abdominal Pain, and PAGI-SYM total), and lastly the Anxiety and Depression sub scores of the PROMIS-29). From this it was able to show two very distinct groupings separating the hEDS from healthy cohorts; however, it should be noted that much of the clustering contribution may be from clear contrasting survey scores. As discussed in detail prior, the immense time to diagnosis and lack of robust diagnostic screening tools for individuals with hEDS is an extremely important gap needing addressed that leads to many harmful outcomes (2,5,11). As is common in complex multisystemic disorders like hEDS, individual parameters likely cannot be used alone in any meaningful medical context due to the high interindividual variability. Our use of exploratory PCA was specifically designed to leverage the interdependencies between metrics and symptoms that could identify a multidimensional phenotype or marker for further investigation into a new screening or monitoring tool. A new screening/diagnostic tool may look similar to how the Apple Watch is currently being used to notify users of potential atrial fibrillation risk, warranting a further check up with a trained professional. Since hEDS is very rare and often overlooked in the initial years of work up in the diagnostic differentials, a simple wearable notification of risk may make a huge difference. Additionally, wearable monitoring could influence clinical decisions with hEDS, if you could identify hEDS patients who have dysautonomia contributing to GI symptoms with these wearable monitors, it could imply treatment for dysautonomia is needed instead of GI medication, or vice versa. It also could provide a continuous monitoring state of the autonomic nervous system while undergoing therapies that affect it (IV lactate ringers, midodrine, electrical nerve stimulation, etc.).

Prior work from Choudhary et. al found that when they stratified a large cohort of EDS individuals (>1000) it was evident that distinct clusters of Disorders of Gut-Brain Interaction (DGBI) exist within the hEDS/HSD population. Specifically, researchers identified two primary groups: those with functional foregut disorders (FFD) overlapping with IBS, and those with predominantly IBS symptoms.

Both clusters demonstrate significantly higher levels of somatic symptoms, autonomic dysfunction, and health-related impairment compared to hEDS/HSD patients without DGBI. Notably, the FFD+IBS subgroup exhibits the most severe clinical profile, with increased rates of anxiety, autonomic symptoms, and comorbid Postural Orthostatic Tachycardia Syndrome (POTS). This prior work may be what we are observing in part with our smaller cohort study, in that it’s possible that our cohort may fall within a specific DGBI cluster they observed, thus likely not generalizable to the larger hEDS population. (50)

Furthermore, while these PCA findings provide some promise for viewing the multidimensional profile of hEDS, they should be interpreted with caution given the relatively modest sample size (n=58). As an exploratory analysis, this study may be susceptible to over-fitting and limited generalizability. The high discriminatory potential identified with PCA should be viewed as a preliminary signal rather than a definitive diagnostic threshold. Regardless, this work warrants further investigation in larger, multicenter cohorts. Furthermore, to use as a screening tool to identify potential patients with hEDS, there likely needs to be a future a control group of non-diabetic gastroparesis patients without joint features of EDS to help show distinguishing capability of non-GI symptoms in the context of the disease. Such expanded studies are necessary to further explore these wearable derived relationships and potential principal component digital biomarkers of hEDS autonomic state to determine if this integrative approach of using wearables and reported symptoms can be developed into a robust screening or monitoring tool for clinical practice.

## CONCLUSION

This work leveraged wearable technology with subjective symptom scores to explore differences in individuals with hEDS and healthy individuals. The findings showed that at a group level, individuals with hEDS take fewer steps than healthy individuals. Individuals with hEDS trends towards higher sympathetic activity and less parasympathetic activity, particularly during sleep. Blood pressure and sympathovagal balance, as evidenced by LFHF ratio, SBP and DBP, are much less stable in the hEDS group compared with healthy controls. Furthermore, within the hEDS cohort, most autonomic wearable metrics are heavily related to their GI symptoms, whereas in healthy controls these metrics relate mainly to psychological symptoms. A combination of key wearable metrics with reported symptoms holds potential for future diagnostic/screening and research tools. In conclusion, this study serves as a foundational step for the use of multi-domain wearable physiological and symptomatic profiling in hEDS with the end goal of improving diagnostic and screening capabilities.

## GRANTS

This study was supported in part by NIH Grants OT2 OD028183-01S3, R01HL158952, and the Indiana University Health-Indiana University School of Medicine Strategic Research Initiative and the Ehlers Danlos Society.

## DISCLAIMERS

The content is solely the responsibility of the authors and does not necessarily represent the official views of Indiana University or Purdue University.

## AUTHOR CONTRIBUTIONS

Ward, Steinhubl, Everett, Nowak, Wo, and Francomano conceived and designed the research. Wilson and Shilling performed the experiment. Wilson analyzed data, interpreted results of experiments, prepared figures, and drafted the manuscript. Wo, Nowak, Everett, Ward and Wilson performed edits and revised the manuscript. Ward approved the last version of manuscript.

## Data Availability

All data produced in the present study are available upon reasonable request to the authors.

